# Human enteric nervous system progenitor transplantation restores functional responses in Hirschsprung Disease patient-derived tissue

**DOI:** 10.1101/2023.11.13.23298455

**Authors:** Benjamin Jevans, Fay Cooper, Yuliia Fatieieva, Antigoni Gogolou, Yi-Ning Kang, Restuadi Restuadi, Pieter Vanden Berghe, Igor Adameyko, Nikhil Thapar, Peter W Andrews, Paolo De Coppi, Anestis Tsakiridis, Conor J McCann

## Abstract

**Objective:** Hirschsprung disease (HSCR) is a severe congenital disorder affecting 1:5000 live births. HSCR results from failure of enteric nervous system (ENS) progenitors to fully colonise the gastrointestinal tract during embryonic development. This leads to aganglionosis in the distal bowel, resulting in disrupted motor activity and impaired peristalsis. Currently, the only viable treatment option is surgical resection of the aganglionic bowel. However, patients frequently suffer debilitating, lifelong symptoms, with multiple surgical procedures often necessary. Hence, alternative treatment options are crucial. An attractive strategy involves the transplantation of ENS progenitors generated from human pluripotent stem cells (hPSCs).

**Design:** ENS progenitors were generated from hPSCs using an accelerated protocol and characterised, in detail, through a combination of single cell RNA-sequencing, protein expression analysis and calcium imaging. We tested ENS progenitors’ capacity to integrate and restore functional responses in HSCR colon, after *ex vivo* transplantation to organotypically cultured patient-derived colonic tissue, using organ bath contractility.

**Results:** We found that our protocol consistently gives rise to high yields of cell populations exhibiting transcriptional and functional hallmarks of early ENS progenitors. Following transplantation, hPSC-derived ENS progenitors integrate, migrate and form neurons within explanted human HSCR colon samples. Importantly, the transplanted HSCR tissue displayed increased basal contractile activity and increased responses to electrical stimulation compared to control tissue.

**Conclusion:** Our findings demonstrate, for the first time, the potential of hPSC-derived ENS progenitors to repopulate and restore functional responses in human HSCR patient colonic tissue.

**What is already known on this topic:**

- Hirschsprung disease is a devastating condition characterized by aganglionosis of the enteric nervous system (ENS) in the distal bowel, leading to dysmotility, severe constipation and enterocolitis.
- Stem cell therapy offers the potential to generate an enteric nervous system in aganglionic tissue and previous studies have described methods for generating ENS progenitors.
- However, the ability of these cells to establish intestinal motility in HSCR human tissue has not been shown.

**What this study adds:**

- We describe, for the first time, the detailed characterization of an ENS progenitor population derived from human pluripotent stem cell lines using our efficient protocol.
- Further, we demonstrate the ability of ENS progenitors to differentiate into enteric neurons *in vitro* and mediate functional rescue following transplantation into explants of human Hirschsprung disease tissue.

**How this study might affect research, practice or policy:**

- These results clearly show the potential of hPSC-derived ENS progenitors in stem cell therapy of Hirschsprung disease for progression towards clinical trials.
- This study highlights the significant advantages of using human surgical discard tissue for testing the efficacy of stem cell therapies.
- The described *ex vivo* model can be used to test different therapeutic approaches prior to clinical trials.

## INTRODUCTION

The enteric nervous system (ENS) is a complex network of neurons and glia organised into ganglia, which controls numerous vital processes in the gut. Defects in ENS generation during embryonic development result in various clinical disorders, which are challenging to treat and associated with considerable morbidity and mortality. The most common enteric neuropathy is Hirschsprung Disease (HSCR), a devastating congenital disease which affects 1:5000 live births and is characterised by the absence of enteric ganglia in the distal portion of the gastrointestinal (GI) tract (1, 2). The aganglionic bowel tonically constricts, causing chronic constipation and a swollen ‘megacolon’ proximal to the aganglionic bowel (3). Children with HSCR are prone to Hirschsprung Disease-associated enterocolitis (HAEC) and bowel perforation (4), which can be fatal if untreated. Surgical resection of the aganglionic region, with pull through of the ganglionic bowel, is currently the only viable treatment option for these children. However, despite surgical intervention, patients often suffer further GI symptoms including HAEC, chronic constipation and intestinal inflammation, with a significant proportion requiring secondary surgeries to resolve ongoing complications (4, 5). Moreover, such complex surgery can lead to long-term consequences such as an unexpectedly high frequency of both subfertility and dyspareunia (6).

HSCR results from a failure in ENS generation. During embryogenesis, the ENS is predominantly derived from a transient population of multipotent cells termed the neural crest (NC) (7, 8). These migrate from the neural tube at the vagal axial level to acquire ENS progenitor features while colonising the GI tract, giving rise to enteric neurons and glia (9–11). In HSCR, ENS progenitors fail to migrate, proliferate or differentiate along variable lengths of the distal gut, which remain aganglionic and fail to function (12). The underlying mechanisms for this failure remain unclear and are likely multifactorial, although disruption of the RET signalling pathway seems to play a key role in a significant number of cases (12–15) A promising strategy for treating HSCR is repopulation of the aganglionic bowel with functional ENS progenitors. Recent proof-of-principle studies have shown unlimited *in vitro* production of human ENS progenitors from human pluripotent stem cells (hPSCs) (16–19). In line with this, we have demonstrated the accelerated and efficient generation of vagal NC cells displaying features of early ENS progenitors from hPSCs (18, 19). These integrated into the mouse GI tract following transplantation and formed ENS-associated neurons and glia (18). However, the full therapeutic potential of these hPSC-derived ENS progenitors remains undetermined. Here, we describe the reproducible generation and detailed characterisation of early ENS progenitors from both embryonic and induced pluripotent stem cells (iPSC). Single cell-RNA sequencing (scRNA-seq) analysis revealed the induction of a population highly expressing vagal NC/early ENS markers, which can be readily directed to differentiate toward functional enteric neurons *in vitro*. Crucially, we also show that hPSC-derived ENS progenitors can integrate and induce positive functional outcomes in patient-derived HSCR colonic tissue following *ex vivo* transplantation. Collectively, these data suggest that human ENS progenitors produced from hPSCs, via our protocol, are a promising cell population for treating HSCR and other enteric neuropathies.

## MATERIALS AND METHODS

### Stem cell culture and differentiation

Use of hESCs has been approved by the Human Embryonic Stem Cell UK Steering Committee (SCSC15-23). The following hPSC lines were employed: WA09/H9 (20), H9-RFP (21), SFCi55-ZsGr (22), MasterShef11 (23) and SOX10:GFP (24). All cell lines were cultured routinely in feeder-free conditions in mTeSR1 (Stem Cell Technologies) medium on Geltrex LDEV-Free reduced growth factor basement membrane matrix (Thermo Fisher). Cells were passaged twice a week after reaching approximately 80% confluency using ReLesR (Stem Cell Technologies) as a passaging reagent. Cells were screened for mycoplasma using Lookout Mycoplasma PCR detection kit (Sigma-Aldrich) or Mycostrip detection kit (Invivogen and were routinely screened for indicators of pluripotency (OCT4, NANOG, **Table S1**) and SSEA4 (25, 26). Neural crest differentiation and enteric neurons were generated as previously described (19) and detailed in the supplementary methods.

### Intracellular staining and flow cytometry

Cells were dissociated using TrypLE Select (Gibco), fixed in Paraformaldehyde (PFA, 4% w/v) for 10 min at room temperature (RT) and permeabilised/blocked with blocking buffer containing 0.1% Triton X-100 and bovine serum albumin (BSA, 1% w/v) in phosphate-buffered saline (PBS) for 1hr at RT. Primary antibodies were diluted in blocking buffer and cells were incubated with anti-SOX10 (**Table S1**) followed by anti-rabbit 488 Alexa fluorophores (A- 21206, Invitrogen). Cells were analysed on a FACS Jazz cell sorter (BD). Data were analysed with FlowJo software (BD).

### Single cell RNA sequencing and analysis

Single cell RNA sequencing was performed by Single Cell Discoveries (Utrecht, The Netherlands), Cells were differentiated as described above. At days 0, 4 and 6 of differentiation, cells were dissociated using TrypLE Select (Gibco) and cryopreserved in STEM-CELLBANKER (AMSBIO). Library preparation was carried out using the 10x Genomics 3’ v3.1 kit followed by sequencing on an Illumina Novaseq 6000. Approximately 10,000 cells per time point were analysed at 30,000 reads/cells, 150 bp, paired end. Detailed RNA sequencing analysis can be found in supplemental methods.

### Calcium imaging

hESC-derived enteric neurons (H9 background) were loaded for 20 min using the Ca^2+^ indicator Fluo-4AM (Invitrogen). Cells were grown on 18mm glass coverslips and transferred to a specifically designed recording chamber. Images were taken using TILLVISION software (TILL Photonics) with a Zeiss Axiovert 200M microscope equipped with a monochromator (Poly V) and a cooled CCD camera (Imago QE), both from TILL Photonics. Fluo-4AM was excited at 470nm and its fluorescence emission collected at 525nm using a 20X (NA 0.75) objective. Images were collected and analysis was performed using custom written macros in IGOR PRO (Wavemetrics). All recordings were performed at RT. To analyse the recorded images Regions Of Interest (ROIs) were drawn over each cell and fluorescence intensity was calculated and normalized per ROI to its baseline starting value. Changes in fluorescence intensity were calculated and expressed as a fraction of the baseline fluorescence, as F/F0. For neuron depolarization, a high K^+^ solution was applied. Cells were also stimulated in an electric field using a 2 second stimulation at 20Hz (40 stim in total) or upon treatment with the nicotinic agonist DMPP (10^-5^M, Sigma Aldrich).

### Processing of patient-derived HSCR colonic surgical discard tissue

Human colonic surgical discard tissue from HSCR patients was obtained with informed consent through Great Ormond Street Hospital, under local REC approval (04/Q0508/79 and 18/EE/0150). Tissue samples were processed under sterile conditions. Briefly, tissue was washed in PBS with Primocin antibiotic (1:500 v/v, InvivoGen) and denuded of the mucosa using fine forceps and micro-scissors. The serosa was removed along with any fatty tissue/blood vessels. From each patient sample, dependent on sample size, up to a maximum of 4 sections of tissue (approx. 1cm^2^, harvested from representative points along the oral-anal axis) were isolated for transplantation alongside an equivalent segment (radially adjacent) as a control for sham transplantation. Transplant and sham tissue samples were pinned in sylgard-coated dishes filled with culture media (DMEM F12 with HEPES and glutamine, supplemented with N2 (Invitrogen), B27 (Invitrogen) and Primocin (500mg/mL, Invivogen)) and placed in a humidified incubator (37°C, 5% CO2) overnight.

### Transplantation of ENS Progenitors to human HSCR colonic tissue

Frozen vials of ENS progenitors were thawed and resuspended in culture media (approx. 125,000 cells/µL). Cell viability and density was assessed using trypan blue dye (Thermo Fisher Scientific Inc.) and an automated cell counter (TC20 Automated Cell Counter, Biorad). Injections into human tissue samples were made using a blunt 5µL Hamilton syringe attached to a pulled glass capillary needle with a tip diameter of 60µm. 4µL cell solution (500,000 cells in total) was injected at a rate of 0.5µl/min. Following injection, the capillary was kept in place for 1 minute before withdrawal. Sham samples received an injection of vehicle-only under the same conditions. Samples were returned to the incubator and maintained for 3 weeks, with media changed every other day.

### Organ bath contractility

Human tissue samples were transferred from culture media to oxygenated Krebs solution and mounted in organ baths (10ml, SI-MB4; World Precision Instruments Ltd). Samples were connected to force transducers (SI-KG20, World Precision Instruments Ltd) via sutures (size 4.0, Fine Science Tools) under an initial tension of 1g. Organ baths were maintained at 37°C and received periodic perfusion of oxygenated Krebs solution. After a 1hr equilibration period contractile activity was recorded using a Lab-Trax-4 data acquisition system (World Precision Instruments Ltd). Tissue samples were subjected to electrical field stimulation (EFS) for 30s (5Hz; 40V; 0.3ms pulse duration) via platinum electrode loops located at both ends of the tissue sample using a MultiStim System (D330, World Precision Instruments Ltd), in the absence or presence of TTX (1µM). Following contractility analysis, samples were harvested for fixation and sectioning.

Contractility data were collected, stored and analysed via Labscribe 2-software (World Precision Instruments Ltd) including analysis of baseline contractile responses (frequency and amplitude) and response to electrical stimulation (Area under the curve (AUC) for the duration of electrical stimulation). To account for tissue variability all raw responses were normalised to wet tissue weight (g).

### Statistics and data presentation

GraphPad Software (GraphPad Prism) was used to generate all graphs and conduct statistical analysis. Data are expressed as mean ± standard error of the mean. The significance of functional data was assessed using a paired Student’s t-test, with p-values <0.05 taken as significant. The individual *ex vivo* tissue samples examined are reported as “segments”. The ‘n values’ reported refer to the number of patients from which tissue samples were harvested. Inter-patient sample pairwise comparisons, between sham-transplanted and ENS progenitor-transplanted human HSCR tissue, were analysed using paired student’s t-tests. To enable analysis across the transplanted cohort (i.e., comparison between responses across all patient tissue samples) and to account for variability between patient samples (age range of 2 months to 4.5 years; 6 males and 1 female) functional contractility data were normalised within individual patient groups using feature scaling (min-max):

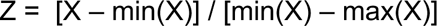

Normalised results (Z) were then analysed using an unpaired Student’s t-test, with p-values <0.05 taken as significant.

Additional methods are provided in online supplementary information.

## RESULTS

### Robust generation of early ENS progenitors from hPSCs

To evaluate the robustness of our previously published vagal NC/ENS differentiation protocol (**Fig. 1A**) (18), we examined its compatibility with two distinct human embryonic (hESC) lines and one induced pluripotent (iPSC) stem cell line. We employed (i) the female hESC line WA09 (H9), including genetically modified versions containing either a constitutive RFP or GFP reporter under the transcriptional control of SOX10 (21, 24), (ii) the male hESC cell line MasterShef11 (MShef11) (23) and (iii) the female iPSC line (SFCi55ZsG) marked by constitutive expression of the fluorescent tag ZsGreen, thus enabling cell tracking and visualisation in transplantation experiments (18, 22). Our protocol initially produces an anterior NC progenitor (ANC) population through combined stimulation of WNT signalling, TGFβ signalling inhibition and moderate BMP activity for 4 days (27), followed by addition of retinoic acid (RA) for a further 2 days (**Fig.1A**). Culture of all 3 cell lines under these conditions efficiently induced the consistent expression of the pan-NC/ENS progenitor marker SOX10 (28, 29) by day 6 of differentiation: 60-80% of all cells were marked by protein expression of SOX10 (**Fig. 1B)**. qRT-PCR analysis of the resulting cultures further demonstrated that the upregulation of *SOX10* was accompanied by high expression levels of *HOX* paralogous group (PG) members indicative of a vagal axial identity (*HOXB4, HOXB5* and *HOXB7*) (**Fig. 1C**). Furthermore, flow cytometry analysis of day 6 cultures derived from H9 background hESCs revealed widespread positivity for the NC/ENS progenitors cell surface markers p75 (NGFR) and CD49d (97.9% and 82.3%, respectively; **Fig. 1D**). Moreover, we detected minimal presence of ‘contaminating’ central nervous system neurectoderm and non-neural ectoderm cells, marked by the expression of PAX6 and GATA3 proteins respectively (<1% of total cells; **Fig. S1A**). Together, these data confirm that our protocol efficiently and rapidly gave rise to high yields of vagal NC/early NC progenitor cells from different hPSC lines.

**Figure 1:**
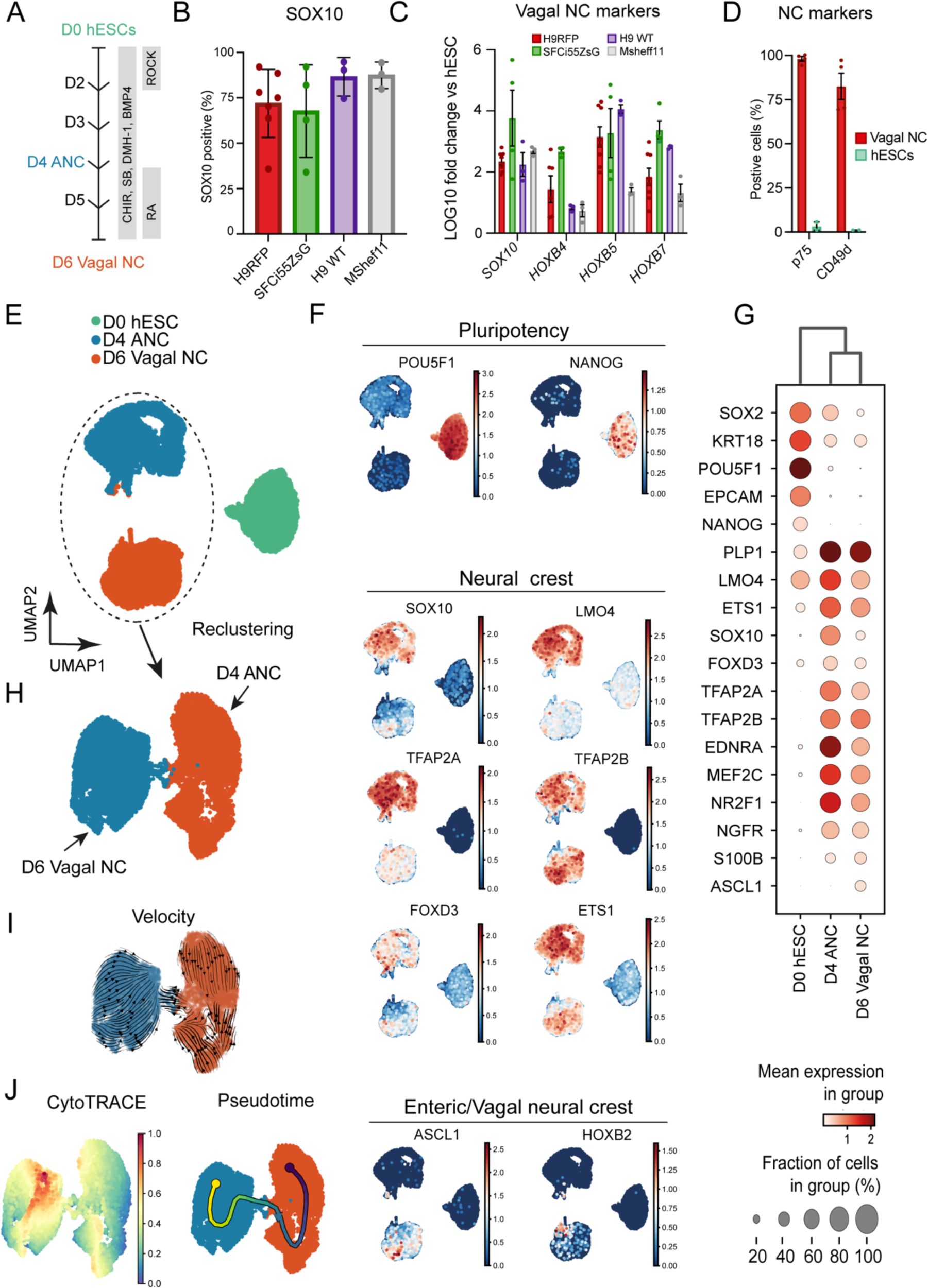
High yields of homogenous vagal NC can be robustly generated from hESCs and iPSCs (A) Schematic representation of the treatment conditions used to generate ENS progenitors *in vitro*. (B) Flow cytometry-based quantification of SOX10 expressing cells in day 6 cultures derived from the indicated hPSC lines. Error bars = SEM (n=3-7 independent differentiations). (C) qPCR expression analysis of *SOX10* and key vagal *HOX* genes carried out in the same D6 cultures as those shown in B (Error bars = SEM; n=3-7 independent differentiations). (D) Quantification of p75 and CD49d-positive cells in day 6 cultures following immunostaining and image analysis (Error Bars = SEM; n=3). Each biological repeat is represented by a unique symbol. (E) UMAP visualization of 17928 cells and their distribution in three samples (D0, D4 and D6) corresponding to unbiasedly defined clusters. (F) UMAP plots showing expression of a set of selected marker genes. (G) Dot plot visualization of the selected expressed marker genes for each cluster. (H) Selective re-clustering of D4 and D6 samples. (I) RNA velocity analysis of D4 and D6 samples (J) Cytotrace and pseudotime analysis of D4 and D6 samples.

We next sought to map the differentiation trajectory of hPSCs toward vagal NC/early ENS progenitors in more detail. To this end, we analysed differentiating hESCs (H9-RFP) at days (D) 0, 4 and 6 by single-cell RNA sequencing (scRNA-seq). We obtained 17,928 cells, which passed quality control (**Fig. S1B-F**), that were allocated to three distinct clusters corresponding to the analysed differentiation timepoints (**Fig. 1E**). The D0 cluster was enriched in pluripotency-associated transcripts (e.g., *POU5F1*, *NANOG*), while the vast majority of cells in the D4 cluster exhibited expression of *bona fide* NC markers such as *SOX10*, *NGFR*, *LMO4*, *ETS1*, *MEF2C* and *TFAP2A/B* (**Fig. 1F&G**). Expression of these transcripts, most of which also are also present in the developing ENS, was largely maintained in the D6 cluster (**Fig. 1F, G**). Cells within the latter uniquely exhibited upregulation of markers specifically denoting a vagal NC/early ENS progenitor state with gliogenic characteristics (PLP1, *ASCL1*, *S100β*, *HOXB2*) (30) (**Fig. 1F, G**). Examination of cell differentiation status was achieved by assigning the developmental stage of cells to clusters connected by trajectory using RNA velocity (31) and also CytoTRACE, a computational framework for predicting differentiation states based on transcriptional diversity (32). This revealed a single stable developmental trajectory from D4 anterior NC cells to D6 vagal NC (**Fig. 1H-J)**. Collectively, these findings confirm that cultures generated using our protocol are predominantly composed of early ENS progenitors; these appear to emerge exclusively upon RA-driven “posteriorisation” of a *HOX*-negative cranial NC state, in line with our previous observations (33).

To assess the capacity of our D6 early ENS progenitors (H9-RFP) to generate enteric-like neurons *in vitro*, we subjected them to ENS differentiation conditions, as previously described (18, 19). By D21, progenitors gave rise to complex neuronal networks widely expressing enteric neuronal (PGP9.5, HUC/D, TH, RET, nNOS) and ENS progenitor/enteric glial markers (SOX10) (**Fig. 2A-L, S2A**). Similar results were obtained using the SFCi55ZsG iPSC line (**Fig. S2B**). Moreover, we found that D6 ENS progenitors could efficiently generate enteric neurons and glia even after cryopreservation and storage in liquid nitrogen for 6 months despite a decrease in viability (**Fig. S2C, D**). The functionality of *in vitro*-derived neurons was further examined via calcium imaging. Ca^2+^ transients in ENS progenitor derivatives were observed following application of either high K^+^, electrical field stimulation or treatment with the nicotinic agonist DMPP (**Fig. 2M-O**). Based on these data, we conclude that D6 hPSC-derived vagal NC/early ENS progenitors can be directed to produce *in vitro* cells that exhibit the hallmarks of functional enteric neurons.

**Figure 2:**
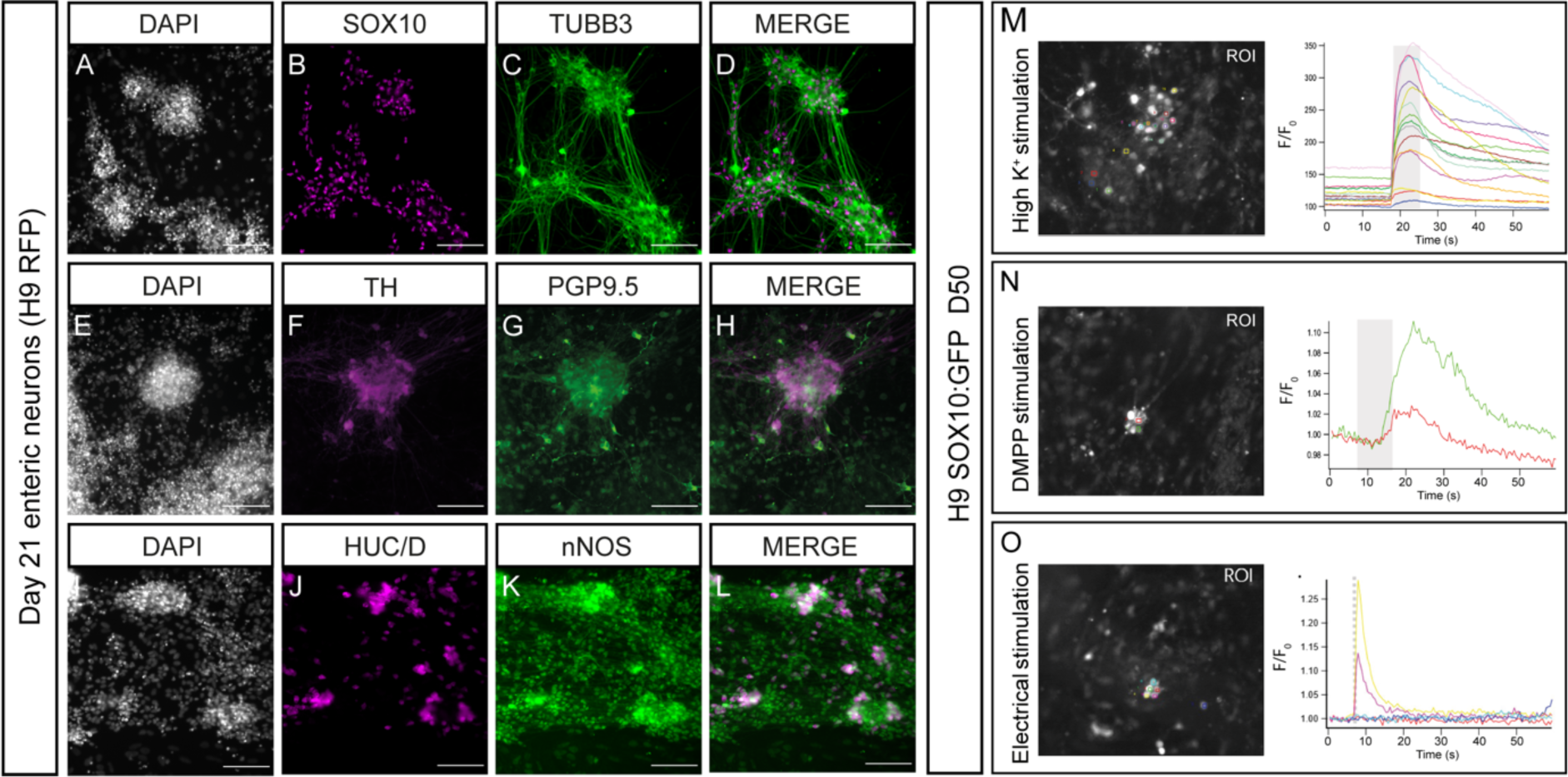
Vagal NC can be differentiated *in vitro* to enteric-like neurons A-L) Representative images of Day 21 enteric neurons immunolabeled with indicated ENS markers following *in vitro* differentiation of hESCs (H9 background). Scale bar = 100µm. (M-O) Analysis and quantification of hESC-derived (H9 background) neuronal Ca^2+^ response to depolarization with high K^+^ (M), activation with DMPP (N) and electrical stimulation (O). Left: overview of the regions of interest (ROI) employed for analysis. Right: individual line traces of the responding cells, the change in fluorescence (F/F0) is plotted over time (s). The traces are randomly chosen for illustration. The application of high K^+^/DMPP/electrical stimulation is represented by the grey bar in the graph.

### hPSC-derived ENS progenitors integrate and form neurons in human HSCR colon following *ex vivo* transplantation

We next tested the ability of our hPSC-derived ENS progenitors to integrate and form neurons in human HSCR colon following *ex vivo* transplantation. HSCR patient-derived colonic samples (obtained from surgical discard tissue under informed consent) were transplanted with freshly thawed vials of frozen ENS progenitors generated from either ZsGreen^+^ SFCi55 iPSCs or H9-RFP hESCs and maintained in organotypic culture *ex vivo* (**Fig. 3A-C**). At the time of transplantation (D0), ZsGreen^+^ ENS progenitors were readily observed at the transplant site (**Fig. 3D).** At D21, ZsGreen expression was maintained and observed in all transplanted segments that were imaged prior to further processing (15/15), with donor cells visualised both at the transplant site and migrating through the surrounding tissue (**Fig. 3E**). Interestingly, donor ENS progenitors appeared to migrate away from the transplant site as individual cells or in “chains” reminiscent of NC cells/ENS progenitors colonising the embryonic murine GI tract *in vivo* (34). These migrating transplanted cells were often detected in close association with the presumptive endogenous tissue vasculature (**Fig. 3F**).

**Figure 3:**
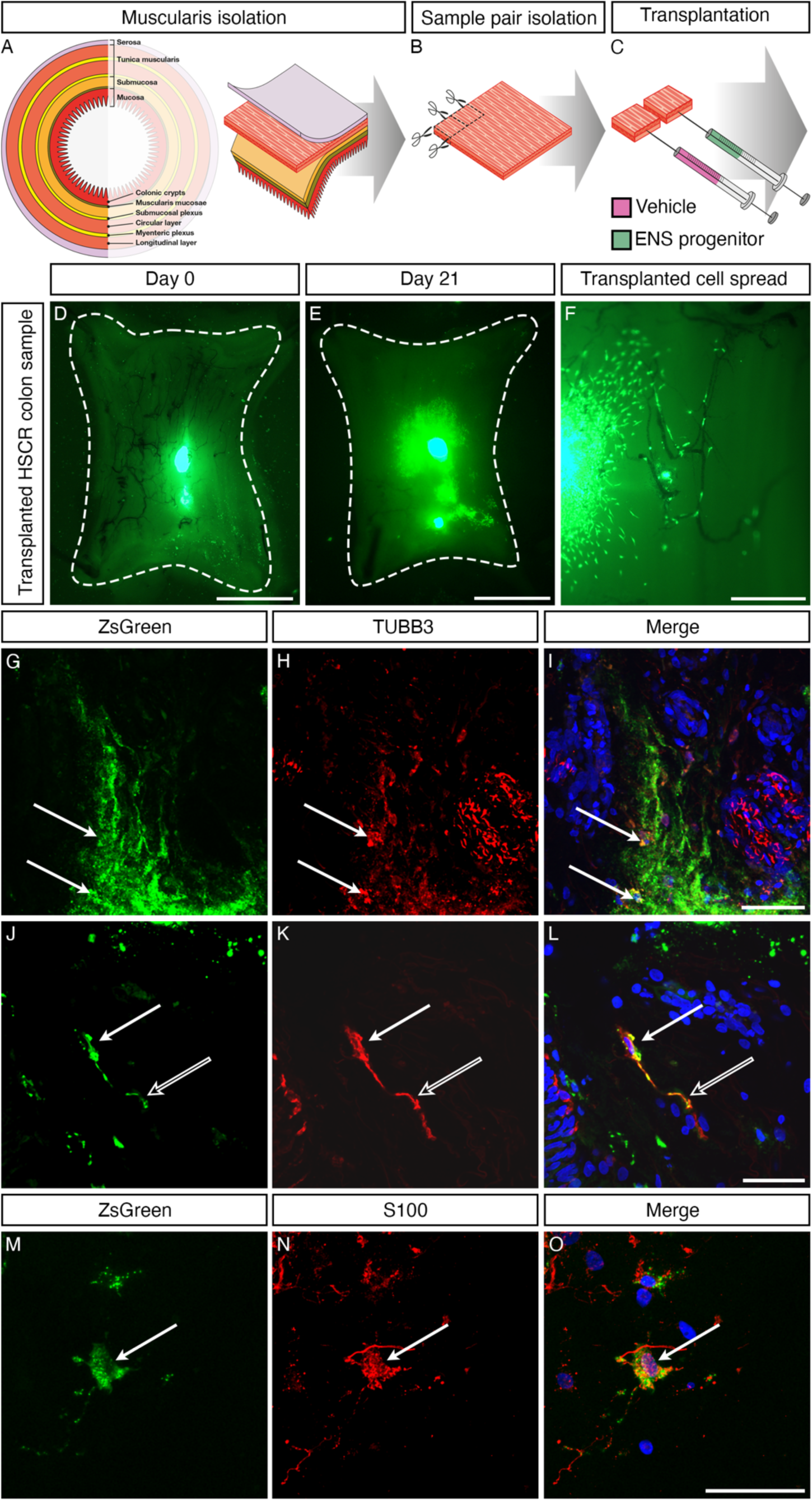
ZsGreen-labelled ENS progenitors survive transplantation into Hirschsprung disease surgical discard tissue, spread and differentiate into neurons (A-C) Schematic representation of *ex vivo* transplantation procedure conducted on patient-derived HSCR colonic tissue. (D-F) Representative stereoscopic images of ZsGreen^+^ donor ENS progenitor cells following *ex vivo* transplantation into patient derived HSCR tissue at day 0 (D0; *D*) and D21 (*E).* High magnification imaging revealed donor cells migrating from the site of transplantation, apparently following blood vessels (*F*). (G-O) Representative confocal images at D21. ZsGreen^+^ ENS progenitors could be seen clustered around the initial transplant site and migrating from the transplant site in streams. Numerous instances of ZsGreen^+^, TUBB3^+^ cells were detected at the presumptive transplant site (*G-I; arrows*). Higher magnification revealed ZsGreen^+^, TUBB3^+^ cells with typical neuronal morphology, including the projection of axon-like processes (*J-L*; hollow arrows). S100+, TUBB3^+^ cells were also detected (*M-O; arrows*). Scale bars: D,E – 2mm; F – 500µm; G-I – 100µm; J-O – 50µm.

To determine whether ENS progenitors transplanted into the HSCR colonic tissue microenvironment retained the neuroglial potential demonstrated *in vitro,* immunohistochemistry was performed for the neuronal marker TUBB3 and glial marker S100. At D21, ZsGreen^+^, TUBB3^+^ donor cells were readily detected within HSCR colonic tissue (**Fig. 3G-I, arrows**) with differentiated donor cells displaying a neuronal morphology, including extension of ZsGreen^+^, TUBB3^+^ axon-like processes (**Fig. 3J-L, hollow arrows**). S100^+^, TUBB3^+^ donor cells were also detected (**Fig. 3M-O, arrows**). Collectively, these results indicate that hPSC-derived ENS progenitors can efficiently migrate, integrate and differentiate within human HSCR colonic tissue *ex vivo*.

### Transplantation of hPSC-derived ENS progenitors restores functional responses in HSCR colonic tissue

The inability of aganglionic colonic tissue to contract and relax in a controlled manner is a key feature of HSCR. As transplanted ENS progenitors were found to integrate and differentiate within the explanted HSCR colon, we next examined their ability to alter the functional responses of the host tissue. Organ bath contractility, performed on transplanted HSCR colonic segments and spatially adjacent sham-injected controls, revealed significant improvements in baseline contractility and response to electrical field simulation (**Fig. 4A-G**). Basal contractile frequency was comparable between ENS progenitor-transplanted segments and sham-transplanted control tissue (10.7 and 16.9 contractions/min, respectively, p=0.2305, **Fig. 4B**, n=7 patients, pairwise comparison of 21 ENS progenitor-transplanted and sham-transplanted segments). Similarly, we detected no difference in basal normalised contractile amplitude between ENS progenitor-transplanted segments and sham-transplanted control tissue (3.2 and 5.2g, respectively, p=0.0887; **Fig. 4C**, n=7 patients, pairwise comparison of 21 paired segments). However, complex basal motor contractions were observed more frequently in hPSC-derived ENS progenitor-transplanted tissue compared to sham-transplanted tissue (61.9% and 42.9%, respectively, n=7 patients, 21 paired segments). This resulted in a significant increase in the cumulative magnitude of basal contractions in ENS progenitor-transplanted tissue compared with sham-transplanted tissue (normalised area under the curve (AUC): 5.8 and 2.7g.s, respectively, p=0.015; **Fig. 4D**, n=7 patients, 21 paired segments). Additionally, ENS progenitor-transplanted tissue elicited a significantly increased contractile response to electrical field stimulation (EFS) when compared to sham-transplanted controls (**Fig. 4E-G**). Normalised contractile amplitude was significantly increased in ENS progenitor-transplanted tissue compared to sham-transplanted controls (9.6 and 4.6g, respectively, p=0.0062; **Fig. 4F**, n=7 patients, 19 paired segments). Similarly, we observed a significant increase in contractile magnitude in ENS progenitor-transplanted tissue compared to sham-transplanted controls (normalised AUC: 157.4 and 68.0g.s, respectively, p=0.0141; **Fig. 4G**, n=7 patients, 19 paired segments). Notably, following application of tetrodotoxin (TTX, 1μM), a potent neurotoxic sodium channel blocker, ENS progenitor-transplanted tissue and sham-transplanted tissue displayed comparable responses to EFS (normalised AUC: 99.5 and 58.6g.s respectively, p=0.0887; **Fig. 4H&I,** n=7 patients, 17 ENS paired segments). We next sought to examine if the observed improvement between paired tissue samples during EFS could be extrapolated at an inter-patient level, taking into account variability in patient age, gender, disease severity and resected tissue size. To achieve this, we employed feature scaling whereby functional responses were normalised across each patient. This revealed a positive increase in the response to EFS in 68.4% of ENS progenitor-transplanted HSCR segments compared to sham-transplanted controls (**Fig. 5A-G**, n=7 patients, 19 paired segments). Promisingly, using this analysis, an overall improvement in the response of ENS progenitor-transplanted segments compared to sham-transplanted controls could be detected at a group level (average slope gradient = +0.3573) which was found to be significant when compared to zero (i.e., no effect, p=0.0108; **Fig. 5H**, n=7 patients). These data suggest an average positive improvement in contractile response to EFS across the entire patient cohort in our study. Collectively, these findings indicate that transplantation of hPSC-derived ENS progenitors triggers neuronally-mediated functional responses in HSCR patient-derived colonic tissue.

**Figure 4:**
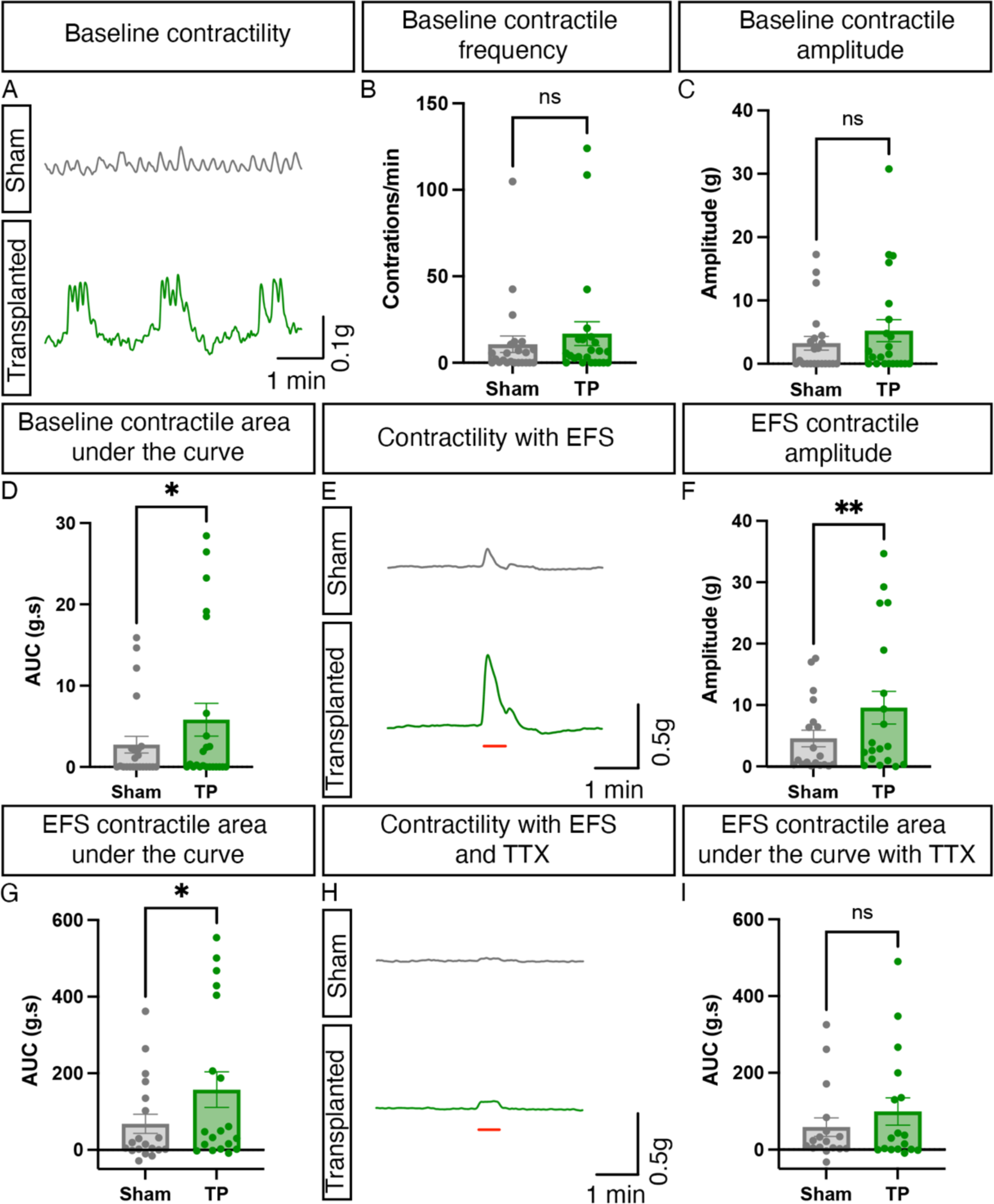
Transplantation of ENS progenitors into Hirschsprung disease surgical discard tissue increases both baseline contractility and response to electrical stimulation via neuronal activity (A) Representative traces showing basal contractile activity in HSCR patient derived tissue 21 days after sham (grey) or ENS progenitor-transplantation (green). (B-D) Summary data of baseline contractile frequency (*B*), amplitude *(C)* and cumulative Area Under the Curve (AUC; *D*). (E) Representative contractility traces of sham (*grey*) or ENS progenitor transplanted (*green*) tissues in response to electrical field stimulation (EFS; *red bar*) in control conditions. (F,G) Summary data of amplitude *(F)* and AUC *(G)* in response to EFS in control conditions. (H) Representative contractility traces of sham- (*grey*) or ENS progenitor-transplanted (*green*) tissues in response to electrical field stimulation (EFS; *red bar*) in the presence of 1μM TTX. (I) Summary data of AUC in response to EFS in TTX. * = p<0.05 Error bars = SEM.

**Figure 5:**
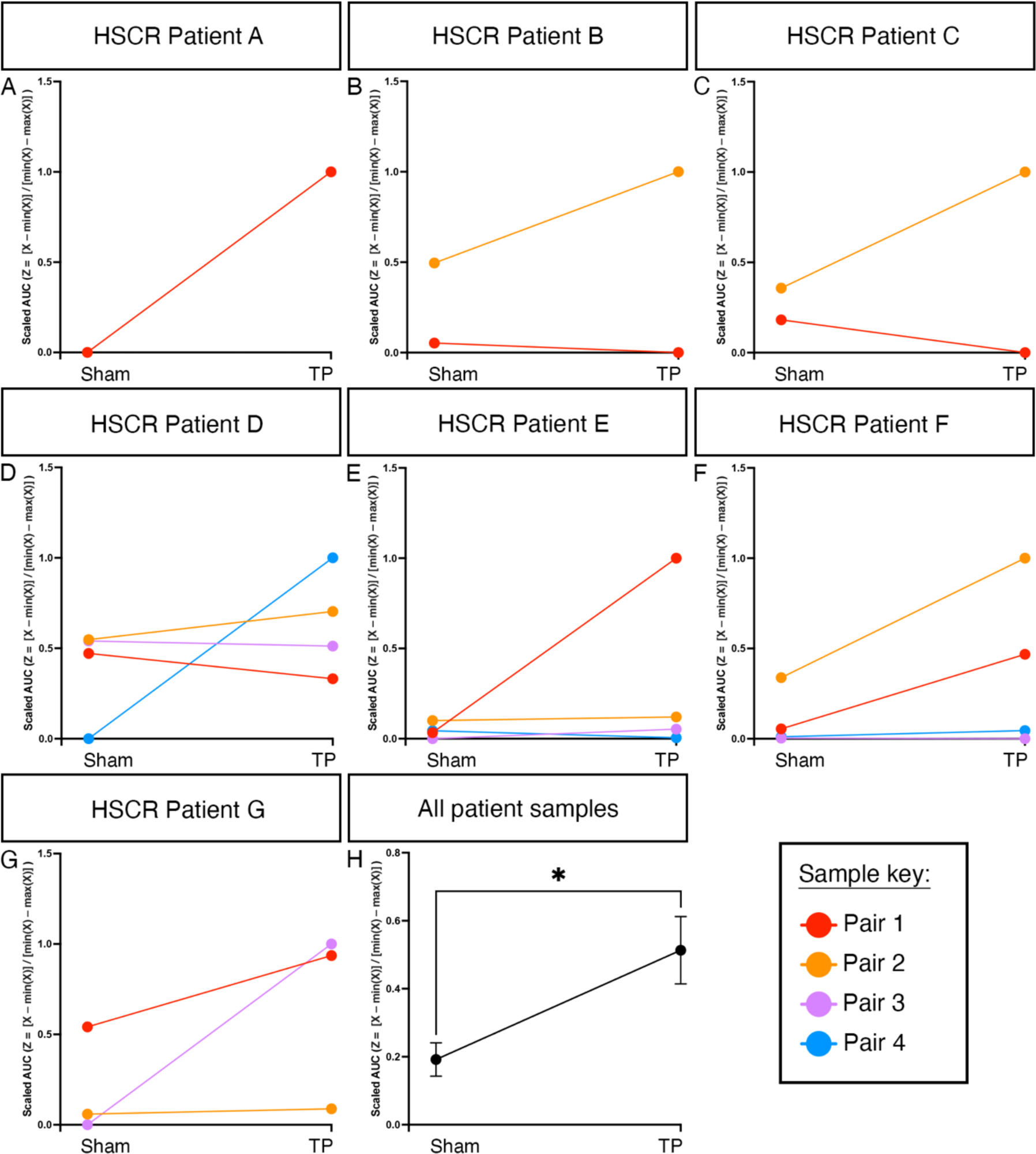
ENS progenitor transplantation increases the response of Hirschsprung disease surgical discard tissue samples to electrical stimulation To account for patient variability (in terms of age, gender, disease severity and positioning of the sample along the oro-anal axis) samples were normalised by patient using feature scaling (Z = [X – min(X)] / [min(X) – max(X)]). (A-G) Slope analysis for each pair of tissue samples under examination by patient. (H) Summary data showing scaled AUC analysis of ENS progenitor-transplanted samples compared to sham-transplanted. * = p<0.05 Error bars = SEM.

## DISCUSSION

HSCR is a debilitating condition with no cure. Although surgery is lifesaving, many HSCR patients suffer persisting symptoms even following surgical intervention (35). Recent advances in stem cell therapy (36, 37) offer the potential to replenish missing neurons following surgery and bridge connections in the newly anastomosed colon to mitigate these ongoing symptoms and possibly, one day, to even remove the need for surgical resection entirely. Previous data have highlighted the potential of autologous (38, 39) and hPSC-derived ENS progenitors (16, 17) to integrate and rescue murine models of colonic dysfunction. Here, we demonstrate that transplantation of ENS progenitors, generated from hPSCs using our previously described protocol (18, 19) and subsequent differentiation *in situ,* as evidenced by detection of donor-derived neurons, can restore functional responses in human HSCR surgical discard tissue. To our knowledge, the results described here represent the first rescue-of-function of human HSCR tissue. We show an increase in the magnitude of basal contractility as well as increased responses to electrical stimulation following ENS progenitor transplantation. Crucially, this finding addresses a key outcome for evaluation of stem cell transplantation efficacy in HSCR, in line with the accepted standards laid out in a ‘white paper’ (40).

We show that our protocol can reproducibly and efficiently give rise to high yields of ENS progenitors from multiple hPSC lines within 6 days. This is quicker than previously published protocols that yield ENS progenitors after 10–15 days at a comparable efficiency (16, 41) thus reducing potential manufacturing costs, a key advantage for the development of a viable cell therapy against HSCR. Our data also indicate that the resulting ENS progenitors can be cryopreserved and recovered with high efficiency which is essential for multicentre clinical application of this technology. Importantly, following recovery, cells maintained their ability to differentiate to ENS lineages and could be utilised for *ex vivo* cell transplantation into

HSCR colonic tissue. Hence, we demonstrate, for the first time, an “off-the-shelf” utility of hPSC-derived ENS progenitors which overcomes a key technical barrier to clinical translation. A relatively modest number of transplanted cells (500,000/cm^2^) was sufficient to promote positive functional change, in line with previous studies showing that small numbers of transplanted cells can exert a dramatic functional effect (42, 43). Although our data indicate that the observed functional rescue is likely to be a result of *de novo* neurogenesis, driven by transplanted ENS progenitors, the possibility of donor cell-mediated non-cell autonomous effects cannot be excluded (38). Our protocol gives rise to cultures that are predominantly (67-87% of total cells) composed of SOX10-positive vagal NC/early ENS progenitors. scRNA-seq analysis of the remaining SOX10-negative fraction indicates that it likely represents more differentiated posterior cranial/vagal NC derivatives and in the future, it will be important to test the effect of the remaining SOX10-negative fraction on the capacity of the transplanted cells to induce functional rescue.

We describe the use of human HSCR patient-derived organotypic tissue. This circumvents some of the issues encountered with HSCR mouse models (e.g., intra and inter model variability in disease severity and short life span), which have slowed efforts to date. Crucially, our organotypic transplantation approach offers a number of advantages which may be useful for future studies including: i) the ability to temporally track transplanted cell integration, ii) the potential to alter cell culture dynamics to affect cell integration/differentiation, iii) an overall reduction in the use of animal-based assays and most importantly iv) the observations achieved through this approach are directly translatable. Remarkably, we were able to achieve extended *ex vivo* human intestinal tissue survival with functional readouts from patient-derived colonic tissue after 3 weeks in culture, providing a powerful preclinical tool for the optimisation of innovative therapies prior to *in vivo* transplantation. However, this model has some technical limitations. While the approach provides a test bed to examine ENS progenitor integration and effects in patient-derived tissue, the relatively small segments (1 cm^2^), used herein, limit understanding of how donor cells may behave in larger specimens. Further, to maintain culture sterility for extended periods it was necessary to remove the mucosal layer prior to transplantation which may impact microenvironmental factors and thus donor cell behaviour. Finally, given the *ex vivo* nature of the model system, signalling components and responses (e.g., immune cells and inflammatory responses) which may influence donor cell efficacy are excluded from our observations.

In conclusion, our findings strongly suggest that hPSC-derived ENS progenitors can serve as the basis for the further development of cell therapies aiming to treat conditions characterised by neuronal loss or dysfunction in the gastrointestinal tract, such as HSCR.

## Data Availability

The data that support the findings of this study are available from the corresponding authors upon reasonable request.

## ACKNOWLEDGEMENTS

The authors thank Dr Dale Moulding (UCL Great Ormond Street Institute of Child Health Imaging Facility) for technical assistance. Part of this research was conducted at UCL Great Ormond Street Institute of Child Health and supported by the NIHR Great Ormond Street Hospital Biomedical Research Centre. Views expressed in this manuscript are solely those of the authors and not necessarily those of the NHS, the NIHR or the Department of Health.

## CONTRIBUTIONS

BJ: conceptualisation, data curation, formal analysis, investigation, methodology validation, visualisation, writing original draft, writing – review editing

FC: conceptualisation, data curation, formal analysis, investigation, methodology validation, visualisation, writing original draft, writing – review editing

YF: data curation, formal analysis, investigation, methodology validation, writing – review editing

AG: data curation, investigation, methodology validation, visualisation, writing – review editing YK: data curation, formal analysis, investigation, methodology validation, writing – review editing

RR: formal analysis, writing – review editing

PVB: conceptualisation, data curation, formal analysis, investigation, writing – review editing IA: data curation, formal analysis, investigation, methodology validation, writing – review editing

NT: conceptualisation, funding acquisition, writing – review editing

PWA: conceptualisation, funding acquisition, project administration, resources, writing – review editing

PDC: conceptualisation, funding acquisition, resources, writing – review editing

AT: conceptualisation, funding acquisition, resources, formal analysis, investigation, supervision, writing original draft, writing – review editing

CMC: conceptualisation, funding acquisition, resources, formal analysis, investigation, supervision, writing original draft, writing – review editing

## SUPPLEMENTARY FIGURE LEGENDS

**Figure S1:**
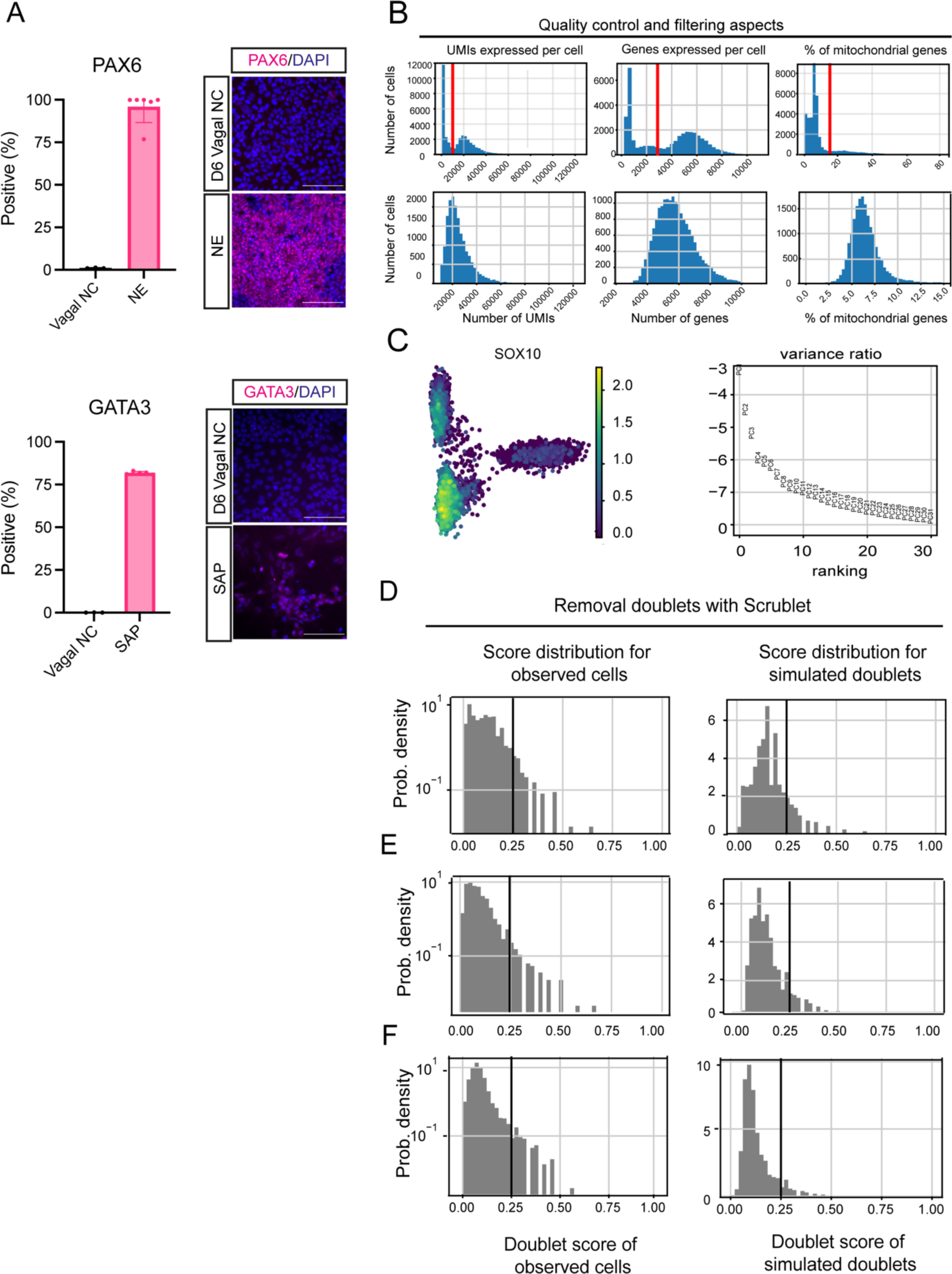
Quality control of single cell RNA sequencing (A) Quantification and representative images of PAX6 and GATA3-positive cells in day (D) 6 neural crest and positive control samples (hPSC-derived neuroectoderm (NE) for PAX6 and sympathetic neurons (SAP) for GATA3) following immunostaining and image analysis. Error Bars = SEM (n=6). Scale bar = 100μm. (B) Plotted number of detected UMIs, genes and percentage of mitochondrial genes per cell before and after filtration. (C) Scatterplot of cells based on the first two principal components and contribution of single PCs to the total variance in the data. (D-F) Application of Scrublet software to D0, D4 and D6 datasets for removing the cell doublets. The left column shows histograms of score distribution for observed cells, the right column shows histograms of score distribution for simulated doublets.

**Figure S2:**
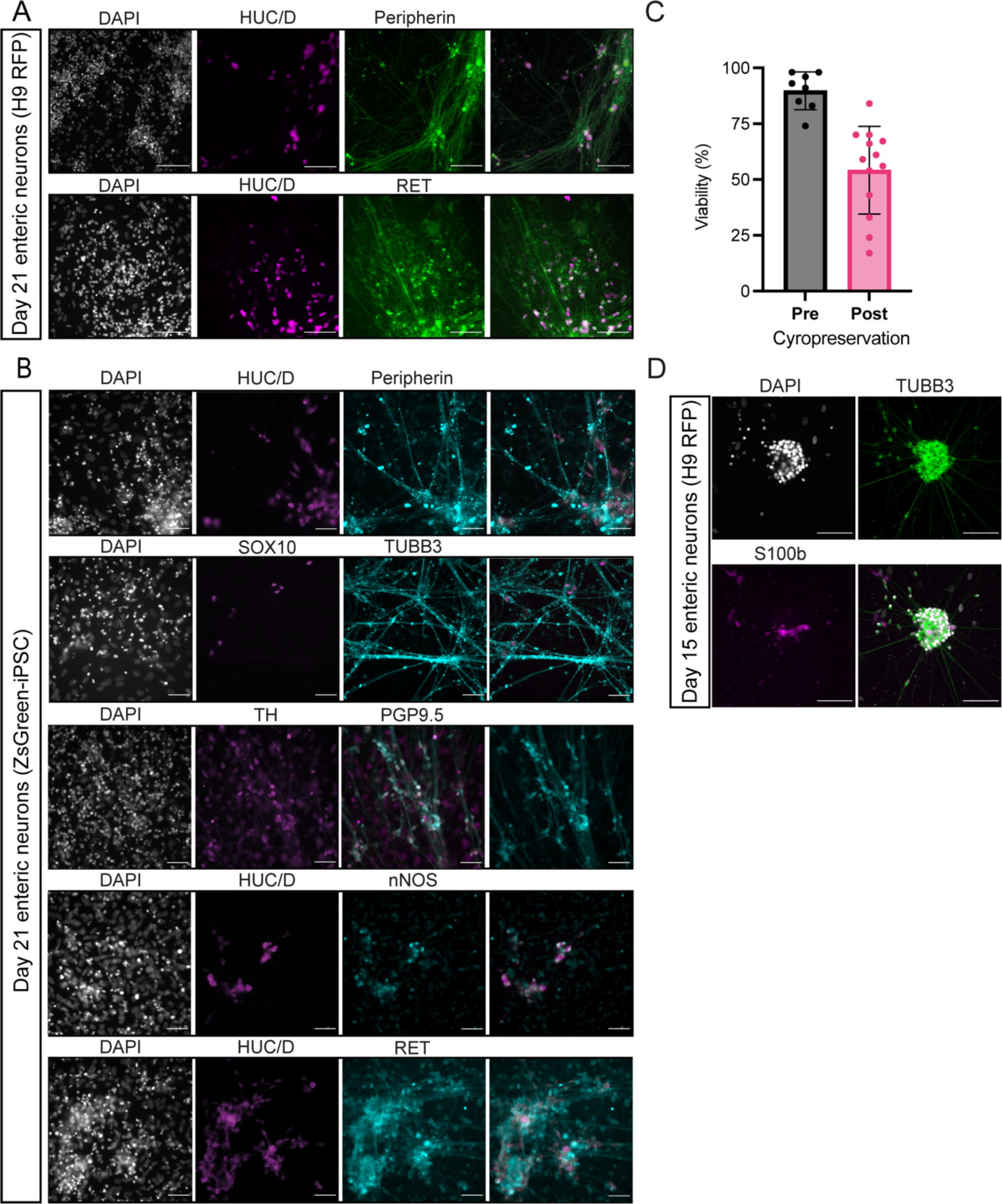
ENS progenitors from hPSCs can be differentiated *in vitro* to enteric-like neurons (A) Representative images of Day (D) 21 enteric neurons immunolabeled with indicated ENS markers following *in vitro* differentiation of H9-RFP. Scale bar = 100μm. (B) Representative images of D21 enteric neurons immunolabeled with indicated ENS markers following *in vitro* differentiation of SFCi55 iPSCs. Scale bar = 100 μm. (C) Viability of ENS progenitors pre- and post- cryopreservation. (D) Representative images of D21 enteric neurons following cryopreservation and storage for 6 months. Neurons are immunolabeled with indicated neural marker (TUBB3) and glia marker (S100B) following *in vitro* differentiation.

## REFERENCES

1. Westfal ML, Goldstein AM. Pediatric enteric neuropathies: diagnosis and current management. Curr Opin Pediatr. 2017;29(3):347–53.

2. Amiel J, Sproat-Emison E, Garcia-Barcelo M, Lantieri F, Burzynski G, Borrego S, et al. Hirschsprung disease, associated syndromes and genetics: a review. Journal of Medical Genetics. 2008;45(1):1–14.

3. Butler Tjaden NE, Trainor PA. The developmental etiology and pathogenesis of Hirschsprung disease. Transl Res. 2013;162(1):1–15.

4. Gershon EM, Rodriguez L, Arbizu RA. Hirschsprung’s disease associated enterocolitis: A comprehensive review. World J Clin Pediatr. 2023;12(3):68–76.

5. Davidson JR, Kyrklund K, Eaton S, Pakarinen MP, Thompson DS, Cross K, et al. Long-term surgical and patient-reported outcomes of Hirschsprung Disease. J Pediatr Surg. 2021;56(9):1502–11.

6. Davidson JR, Kyrklund K, Eaton S, Pakarinen MP, Thompson DS, Cross K, et al. Sexual function, quality of life, and fertility in women who had surgery for neonatal Hirschsprung’s disease. Br J Surg. 2021;108(2):e79–e80.

7. Simões-Costa M, Bronner ME. Establishing neural crest identity: a gene regulatory recipe. Development. 2015;142(2):242–57.

8. Rothstein M, Bhattacharya D, Simoes-Costa M. The molecular basis of neural crest axial identity. Developmental Biology. 2018;444:S170–S80.

9. Sasselli V, Pachnis V, Burns AJ. The enteric nervous system. Dev Biol. 2012;366(1):64–73.

10. Kang Y-N, Fung C, Vanden Berghe P. Gut innervation and enteric nervous system development: a spatial, temporal and molecular tour de force. Development. 2021;148(3).

11. Hutchins EJ, Kunttas E, Piacentino ML, Howard AGAt, Bronner ME, Uribe RA. Migration and diversification of the vagal neural crest. Dev Biol. 2018;444 Suppl 1(Suppl 1):S98-s109.

12. Obermayr F, Hotta R, Enomoto H, Young HM. Development and developmental disorders of the enteric nervous system. Nat Rev Gastroenterol Hepatol. 2013;10(1):43–57.

13. Edery P, Lyonnet S, Mulligan LM, Pelet A, Dow E, Abel L, et al. Mutations of the RET proto-oncogene in Hirschsprung’s disease. Nature. 1994;367(6461):378-80.

14. Tomuschat C, Puri P. RET gene is a major risk factor for Hirschsprung’s disease: a meta-analysis. Pediatr Surg Int. 2015;31(8):701–10.

15. Lyonnet S, Bolino A, Pelet A, Abel L, Nihoul-Fékété C, Briard ML, et al. A gene for Hirschsprung disease maps to the proximal long arm of chromosome 10. Nat Genet. 1993;4(4):346–50.

16. Fattahi F, Steinbeck JA, Kriks S, Tchieu J, Zimmer B, Kishinevsky S, et al. Deriving human ENS lineages for cell therapy and drug discovery in Hirschsprung disease. Nature. 2016;531(7592):105-9.

17. Fan Y, Hackland J, Baggiolini A, Hung LY, Zhao H, Zumbo P, et al. hPSC-derived sacral neural crest enables rescue in a severe model of Hirschsprung’s disease. Cell Stem Cell. 2023;30(3):264–82.e9.

18. Frith TJR, Gogolou A, Hackland JOS, Hewitt ZA, Moore HD, Barbaric I, et al. Retinoic Acid Accelerates the Specification of Enteric Neural Progenitors from In-Vitro-Derived Neural Crest. Stem Cell Reports. 2020;15(3):557–65.

19. Gogolou A, Frith TJR, Tsakiridis A. Generating Enteric Nervous System Progenitors from Human Pluripotent Stem Cells. Current Protocols. 2021;1(6):e137.

20. Thomson JA, Itskovitz-Eldor J, Shapiro SS, Waknitz MA, Swiergiel JJ, Marshall VS, et al. Embryonic stem cell lines derived from human blastocysts. Science. 1998;282(5391):1145-7.

21. Cooper F, Souilhol C, Haston S, Gray S, Boswell K, Gogolou A, et al. Notch signalling influences cell fate decisions and HOX gene induction in axial progenitors. bioRxiv. 2023:2023.06.16.545269.

22. Lopez-Yrigoyen M, Fidanza A, Cassetta L, Axton RA, Taylor AH, Meseguer-Ripolles J, et al. A human iPSC line capable of differentiating into functional macrophages expressing ZsGreen: a tool for the study and in vivo tracking of therapeutic cells. Philos Trans R Soc Lond B Biol Sci. 2018;373(1750).

23. Canham MA, Van Deusen A, Brison DR, De Sousa PA, Downie J, Devito L, et al. The Molecular Karyotype of 25 Clinical-Grade Human Embryonic Stem Cell Lines. Sci Rep. 2015;5:17258.

24. Chambers SM, Qi Y, Mica Y, Lee G, Zhang XJ, Niu L, et al. Combined small-molecule inhibition accelerates developmental timing and converts human pluripotent stem cells into nociceptors. Nat Biotechnol. 2012;30(7):715–20.

25. Adewumi O, Aflatoonian B, Ahrlund-Richter L, Amit M, Andrews PW, Beighton G, et al. Characterization of human embryonic stem cell lines by the International Stem Cell Initiative. Nat Biotechnol. 2007;25(7):803–16.

26. Draper JS, Pigott C, Thomson JA, Andrews PW. Surface antigens of human embryonic stem cells: changes upon differentiation in culture. J Anat. 2002;200(Pt 3):249–58.

27. Hackland JOS, Frith TJR, Thompson O, Marin Navarro A, Garcia-Castro MI, Unger C, et al. Top-Down Inhibition of BMP Signaling Enables Robust Induction of hPSCs Into Neural Crest in Fully Defined, Xeno-free Conditions. Stem Cell Reports. 2017;9(4):1043–52.

28. Bondurand N, Natarajan D, Thapar N, Atkins C, Pachnis V. Neuron and glia generating progenitors of the mammalian enteric nervous system isolated from foetal and postnatal gut cultures. Development. 2003;130(25):6387–400.

29. Kelsh RN. Sorting out Sox10 functions in neural crest development. Bioessays. 2006;28(8):788–98.

30. Memic F, Knoflach V, Sadler R, Tegerstedt G, Sundström E, Guillemot F, et al. Ascl1 Is Required for the Development of Specific Neuronal Subtypes in the Enteric Nervous System. J Neurosci. 2016;36(15):4339–50.

31. Bergen V, Lange M, Peidli S, Wolf FA, Theis FJ. Generalizing RNA velocity to transient cell states through dynamical modeling. Nat Biotechnol. 2020;38(12):1408–14.

32. Gulati GS, Sikandar SS, Wesche DJ, Manjunath A, Bharadwaj A, Berger MJ, et al. Single-cell transcriptional diversity is a hallmark of developmental potential. Science. 2020;367(6476):405-11.

33. Frith TJR, Granata I, Wind M, Stout E, Thompson O, Neumann K, et al. Human axial progenitors generate trunk neural crest cells in vitro. eLife. 2018;7:e35786.

34. Young HM, Bergner AJ, Simpson MJ, McKeown SJ, Hao MM, Anderson CR, et al. Colonizing while migrating: how do individual enteric neural crest cells behave? BMC Biol. 2014;12:23.

35. Widyasari A, Pavitasari WA, Dwihantoro A, Gunadi. Functional outcomes in Hirschsprung disease patients after transabdominal Soave and Duhamel procedures. BMC Gastroenterol. 2018;18(1):56.

36. Hallett PJ, Deleidi M, Astradsson A, Smith GA, Cooper O, Osborn TM, et al. Successful function of autologous iPSC-derived dopamine neurons following transplantation in a non-human primate model of Parkinson’s disease. Cell Stem Cell. 2015;16(3):269–74.

37. Schwartz SD, Regillo CD, Lam BL, Eliott D, Rosenfeld PJ, Gregori NZ, et al. Human embryonic stem cell-derived retinal pigment epithelium in patients with age-related macular degeneration and Stargardt’s macular dystrophy: follow-up of two open-label phase 1/2 studies. Lancet. 2015;385(9967):509-16.

38. McCann CJ, Cooper JE, Natarajan D, Jevans B, Burnett LE, Burns AJ, et al. Transplantation of enteric nervous system stem cells rescues nitric oxide synthase deficient mouse colon. Nat Commun. 2017;8:15937.

39. Hotta R, Stamp LA, Foong JP, McConnell SN, Bergner AJ, Anderson RB, et al. Transplanted progenitors generate functional enteric neurons in the postnatal colon. J Clin Invest. 2013;123(3):1182–91.

40. Burns AJ, Goldstein AM, Newgreen DF, Stamp L, Schäfer KH, Metzger M, et al. White paper on guidelines concerning enteric nervous system stem cell therapy for enteric neuropathies. Dev Biol. 2016;417(2):229–51.

41. Workman MJ, Mahe MM, Trisno S, Poling HM, Watson CL, Sundaram N, et al. Engineered human pluripotent-stem-cell-derived intestinal tissues with a functional enteric nervous system. Nat Med. 2017;23(1):49–59.

42. Pan W, Rahman AA, Stavely R, Bhave S, Guyer R, Omer M, et al. Schwann Cells in the Aganglionic Colon of Hirschsprung Disease Can Generate Neurons for Regenerative Therapy. Stem Cells Transl Med. 2022;11(12):1232–44.

43. Zhou Y, Besner G. Transplantation of amniotic fluid-derived neural stem cells as a potential novel therapy for Hirschsprung’s disease. J Pediatr Surg. 2016;51(1):87–91.

